# Evaluating Performance of Near-Term Influenza Forecasters Across Consecutive Seasons

**DOI:** 10.1101/2023.09.28.23296216

**Authors:** VP Nagraj, Desiree Williams, Stephen D. Turner

## Abstract

Near-term forecasting efforts for seasonal influenza (flu) aim to enable better public health preparedness before, during, and after each season. The FluSight initiative has fostered flu forecasting activities since 2013. In 2021-22, the organizers switched the primary target to incident weekly flu hospitalizations at state, territorial, and national levels in the United States. Here we studied the performance of contributors who submitted forecasts for this target during the 2021-22 and 2022-23 seasons. We found that forecasters generally did not perform consistently across locations within seasons. For the select group of forecasters who submitted to both seasons, the overall performance relative to one another was not always consistent either. However, several forecasters, including the FluSight ensemble, were among the top performers in both seasons.

## 1 Introduction

Seasonal influenza (flu) imposes a significant and ongoing burden on population health around the world (CDC 2022). The severity and strain on public health resources varies by season. To help better anticipate geographic spread and intensity of flu activity, infectious disease modelers have developed near-term flu forecasting methodologies. Forecasters use a variety of computational modeling approaches and data sources from which they can engineer features and estimate parameters for models (Kandula and Shaman 2019; Lu and Meyer 2020; McAndrew and Reich 2021; Osthus 2022). Once disseminated, near-term forecasts provide public health officials and the general public with valuable information to inform resource allocation, vaccination campaigns, and the implementation of preventive measures before, during, and after the flu season.

In recent years, such forecasting activities have become centralized through consortia efforts. Contributors using disparate modeling methodology and data sources are openly invited to contribute forecasts in a common format for eventual ensembling and dissemination. The Centers for Disease Control and Prevention (CDC) FluSight initiative, established in 2013, has been foundational in formalizing the “forecast hub” approach (Reich et al. 2019). The COVID-19 pandemic inspired similar forecasting hubs (Cramer et al. 2022), and also shifted the focus of FluSight. COVID-19 has changed dynamics of influenza-like illness (ILI) (Zipfel, Colizza, and Bansal 2021), which historically was the flu activity indicator used as a target for FluSight forecasting. Likewise, the COVID-19 pandemic motivated data collection efforts that have standardized flu data alongside COVID-19 reporting. One example is the mandatory state-level reporting of flu hospitalizations alongside COVID-19 hospitalization data via the HHS Protect system during the COVID-19 public health emergency. As of the 2021-22 season, the FluSight coordinators leveraged HHS Protect reporting to shift the forecasting target to incident weekly hospitalizations by state for 1-4 week-ahead horizons. The target has remained the same during the 2022-23 FluSight season. The FluSight network includes contributors who have submitted across multiple seasons, in some cases using the same methodology.

Here we primarily aim to assess how forecasters submitting to both the 2021-22 and 2022-23 FluSight seasons performed. We estimate forecaster performance within and between 2021-22 and 2022-23 seasons. Seasonal dynamics of flu can vary dramatically, as illustrated by the 2021-22 season that peaked later compared to previous years (CDC 2023c). Such an analysis will demonstrate if and to what extent performance varies not only across forecast dates, horizons, and locations, but also across seasons. The results of this analysis may help forecasters and hub coordinators understand sensitivity of contributions to seasonal dynamics, and to reinforce the utility of consortium ensemble approaches that have previously been shown to improve forecasting accuracy (Wang et al. 2022; Wu and Levinson 2021).

## 2 Methods

### 2.1 Data sources

#### 2.1.1 Truth data

Daily laboratory-confirmed flu cases among hospitalized patients as reported by hospital systems to the U.S. Department of Health & Human Services HHS Protect system (HHS 2023) were used as the “gold standard” data for evaluation of forecast performance. These data are aggregated to the state level in HHS Protect and further aggregated from daily to weekly resolution to compare against weekly forecasts of incidence. All evaluations compared submitted forecasts to data that was current as of July 2023 in the HHS Protect system.

#### 2.1.2 Forecasts

To evaluate performance we first needed to retrieve published forecast data from FluSight. We retrieved the openly licensed submissions using the Zoltar forecast repository (Reich et al. 2021). The FluSight coordinators allow teams to make multiple submissions for different methods. As such, we consider individual submissions as “forecasters” (i.e., unique combination of team and method). Every weekly forecast submission required both a point estimate and a probabilistic distribution described in 23 quantiles^1^, and forecasts ranged from January 10, 2022 to June 20, 2022 (24 weeks) and October 17, 2022 to May 15, 2023 (31 weeks) for the 2021-22 and 2022-23 seasons respectively. In total, there were 56 contributed forecasters, with 25 in the 2021-22 season and 31 in the 2022-23 season. From these we analyzed a subset of forecasters that regularly submitted within each respective season. For a forecaster to be included, it must contain all quantiles and horizons for each given location and forecast week. Furthermore, for any given forecast week to be included in evaluation we required that the forecasters must include forecasts for at least 25 locations (i.e., approximately 50% of the geographic locations across state, territorial, and national resolution). Finally, across the season the forecaster must have submitted to at least 60% of the forecast weeks. In addition to contributed forecasts, we retrieved the FluSight ensemble forecasts and FluSight baseline forecasts, which were both available for all locations and forecast weeks across the 2021-22 and 2022-23 seasons.

FluSight forecasters are free to use any methods they choose to generate forecasts for submissions. The consortium asks that contributors include a description of methods in a metadata file. We reviewed metadata files for submitting forecasters to ascertain the kinds of methods being used per metadata descriptions. Contributors used a wide variety methods, including deep learning models, regression techniques, various compartmental models, time series approaches, and ensembles to generate forecasts. The FluSight baseline model was developed as a comparison for evaluation of submissions. This model predicts the incidence as equal to value of prior week (CDC 2023a).The FluSight ensemble uses all eligible submitted forecasts^2^ to create a combined forecast for respective targets (CDC 2023b). The FluSight baseline and ensemble approaches remained the same in both the 2021-2022 seasons. For contributing forecasters who submitted to both seasons, we reviewed the history of the metadata to see if there were any significant changes in methods described for 2021-22 versus 2022-23 submissions.

### 2.2 Performance evaluation

To evaluate forecasting performance we use two measures: weighted interval score (WIS) and absolute error (AE). The WIS has been described previously (Bracher et al. 2021b) and is frequently used in contemporary evaluations of infectious disease forecasting performance (Bracher et al. 2021a; Sherratt et al. 2023). While the WIS compares the forecast distribution (i.e., all quantiles), the AE measures the difference between the forecasted point estimate and the observed value. Because the WIS measure considers the entire distribution, it has the advantage of being able to estimate over/under prediction. In short, the WIS assigns weights to different spaces in the prediction interval based on the actual outcome. The WIS is always non-negative, with a lower score indicating better forecast accuracy (smaller error). Likewise, a smaller AE indicates better performance. The AE provides an absolute measure that complements WIS. It may be informative to investigate discrepancies between AE and WIS performance. For example, forecasters that produce accurate point estimates but perhaps have less well calibrated error estimation might exhibit such discrepancy. Therefore we calculated both the WIS and AE using the evalcast R package (McDonald et al. 2023) for every forecaster, forecast week, location, and horizon. To standardize across forecasters we computed a relative WIS (rWIS) compared to the FluSight baseline WIS. We further used medians of rWIS and AE across all forecast weeks, locations, and horizons to rank performance of forecasters relative to one another.

## 3 Results

### 3.1 Hospitalization data

The pattern of incident flu hospitalizations reported in HHS Protect was strikingly different between the 2021-22 and 2022-23 seasons. Fig. 1 displays a heatmap of standardized hospitalization rates across states in each season. The observed peak in 2021-22 was shifted much later than a typical season, with some states seeing the highest rate of hospitalizations as late as May 2022. The 2022-23 season conformed to typical historical trends for flu activity, with peaks in most locations in the winter months (Lowen and Steel 2014; Reichert et al. 2004). However, some locations in the south (e.g., Louisiana, Mississippi, and Alabama) experienced elevated rates as early as October 2022.

**Figure 1:**
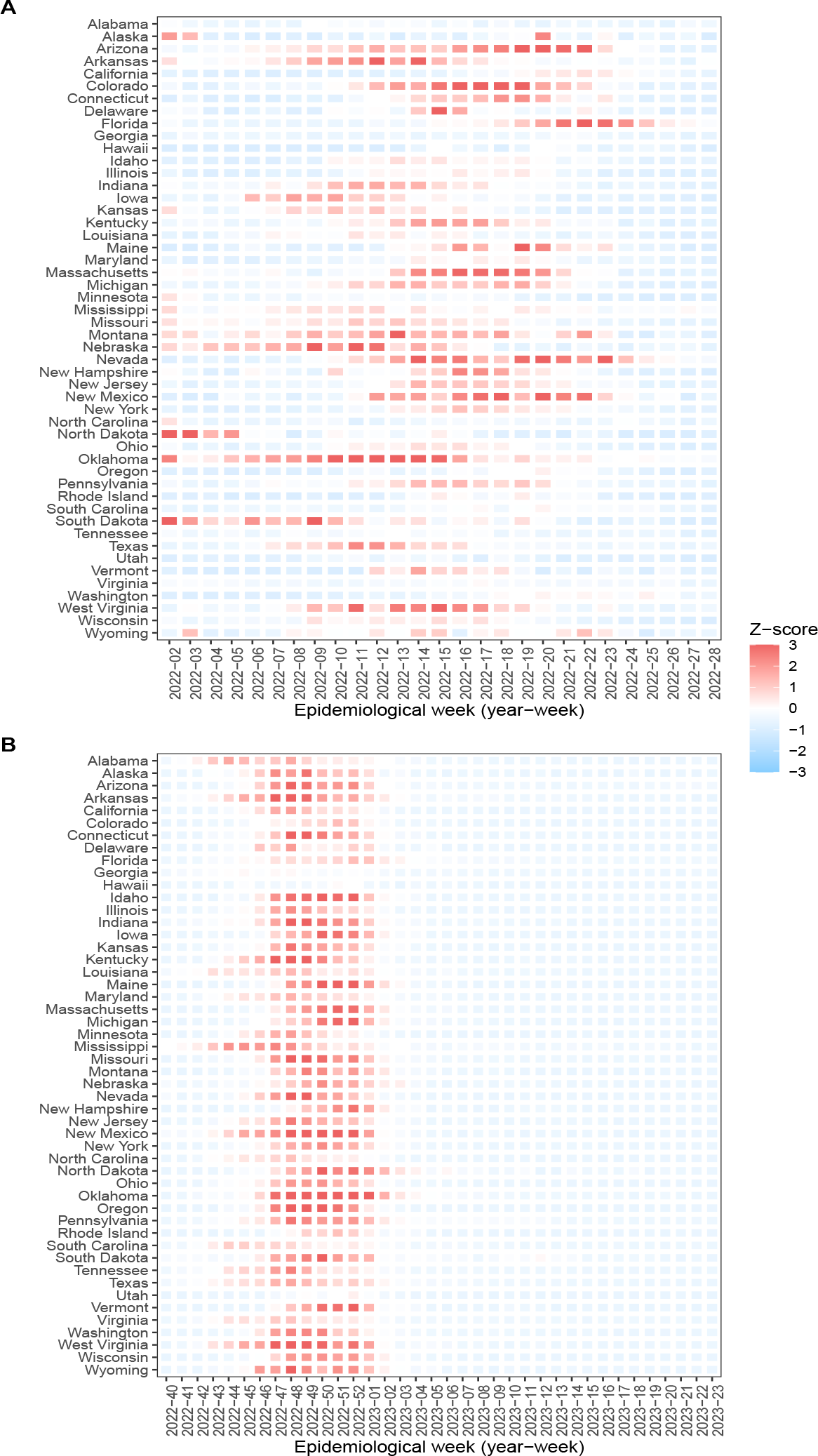
Comparison of the observed hospitalizations across all states. (A) The observed hospitalizations in the 2021-22 flu season. (B) The observed hospitalizations in the 2022-23 flu season. All hospitalizaton counts are converted to a rate per 100,000 based on population of the given state and further standardized with a Z-score within each season. The 2021-22 flu season is truncated to begin in January 2022 to align with initation of flu hospitalization data reporting requirements in HHS Protect, and extends to June 2022 given extended CDC monitoring due to late-season activity.

### 3.2 Forecasters included

Based on our inclusion criteria for completeness of submissions, we excluded three forecasters from the 2021-22 season and 13 from the 2022-23 season. Additionally, one forecaster distributed forecasts with an ambiguous license and was therefore excluded from both seasons. In total, we analyzed 21 contributed forecasters for 2021-22 and 17 for 2022-23. Of these forecasters, 11 submitted to both seasons. The full list of contributing forecasters along with number of weekly submissions and unique locations by season is provided in Table 1. Nearly all of the forecasters submitted all weeks and all locations.

**Table 1:** Summary of forecast submissions by season. The table includes the number of weekly submissions alongside the number of unique locations submitted during the season. Forecasters that submitted to only one season have an ‘X’ in the column for the other season.

|  | Season 2021-2022 | Season 2022-2023 |
| --- | --- | --- |
| Forecaster-01 | 24 weeks; 51 locations | 31 weeks; 51 locations |
| Forecaster-02 | X | 30 weeks; 52 locations |
| Forecaster-03 | X | 25 weeks; 52 locations |
| Forecaster-04 | 24 weeks; 52 locations | 29 weeks; 52 locations |
| Forecaster-05 | 18 weeks; 51 locations | 25 weeks; 52 locations |
| Forecaster-06 | 24 weeks; 52 locations | X |
| Forecaster-07 | 24 weeks; 52 locations | 23 weeks; 52 locations |
| Forecaster-08 | X | 29 weeks; 52 locations |
| Forecaster-09 | 24 weeks; 52 locations | 31 weeks; 52 locations |
| Forecaster-10 | 20 weeks; 52 locations | 30 weeks; 52 locations |
| Forecaster-11 | 24 weeks; 52 locations | X |
| Forecaster-12 | 24 weeks; 51 locations | 31 weeks; 51 locations |
| Forecaster-13 | 22 weeks; 52 locations | X |
| Forecaster-14 | 23 weeks; 52 locations | X |
| Forecaster-15 | 22 weeks; 52 locations | X |
| Forecaster-16 | 22 weeks; 52 locations | X |
| Forecaster-17 | 23 weeks; 52 locations | X |
| Forecaster-18 | X | 28 weeks; 52 locations |
| Forecaster-19 | X | 20 weeks; 52 locations |
| Forecaster-20 | 21 weeks; 52 locations | X |
| Forecaster-21 | 24 weeks; 52 locations | 31 weeks; 52 locations |
| Forecaster-22 | 24 weeks; 52 locations | 26 weeks; 52 locations |
| Forecaster-23 | 21 weeks; 52 locations | X |
| Forecaster-24 | 24 weeks; 51 locations | 29 weeks; 51 locations |
| Forecaster-25 | 22 weeks; 52 locations | 31 weeks; 52 locations |
| Forecaster-26 | 24 weeks; 52 locations | X |
| Forecaster-27 | X | 26 weeks; 52 locations |

### 3.3 Performance across locations

Within each season, we observed that the FluSight ensemble was consistently one of the best performers per the rWIS metric. For the ensemble and individual forecasters, the performance varied by location. In the 2021-22 season (Fig. 2A), several forecasters performed similar to the ensemble, scoring better than baseline on median across most locations. However, most of the forecasters performed no better or worse than baseline on median. Notably, some of the forecasters who scored poorly in some locations scored very highly in others. For example, Forecaster-07 and Forecaster-12 both had the best median rWIS for Kansas and North Dakota respectively but did not perform as well in other locations. It is also worth noting that while for some locations (like Alabama and Wyoming) nearly all of the forecasts beat the baseline, for other locations (like Arizona and Hawaii) there were only one or two forecasters with a median rWIS < 1.

**Figure 2:**
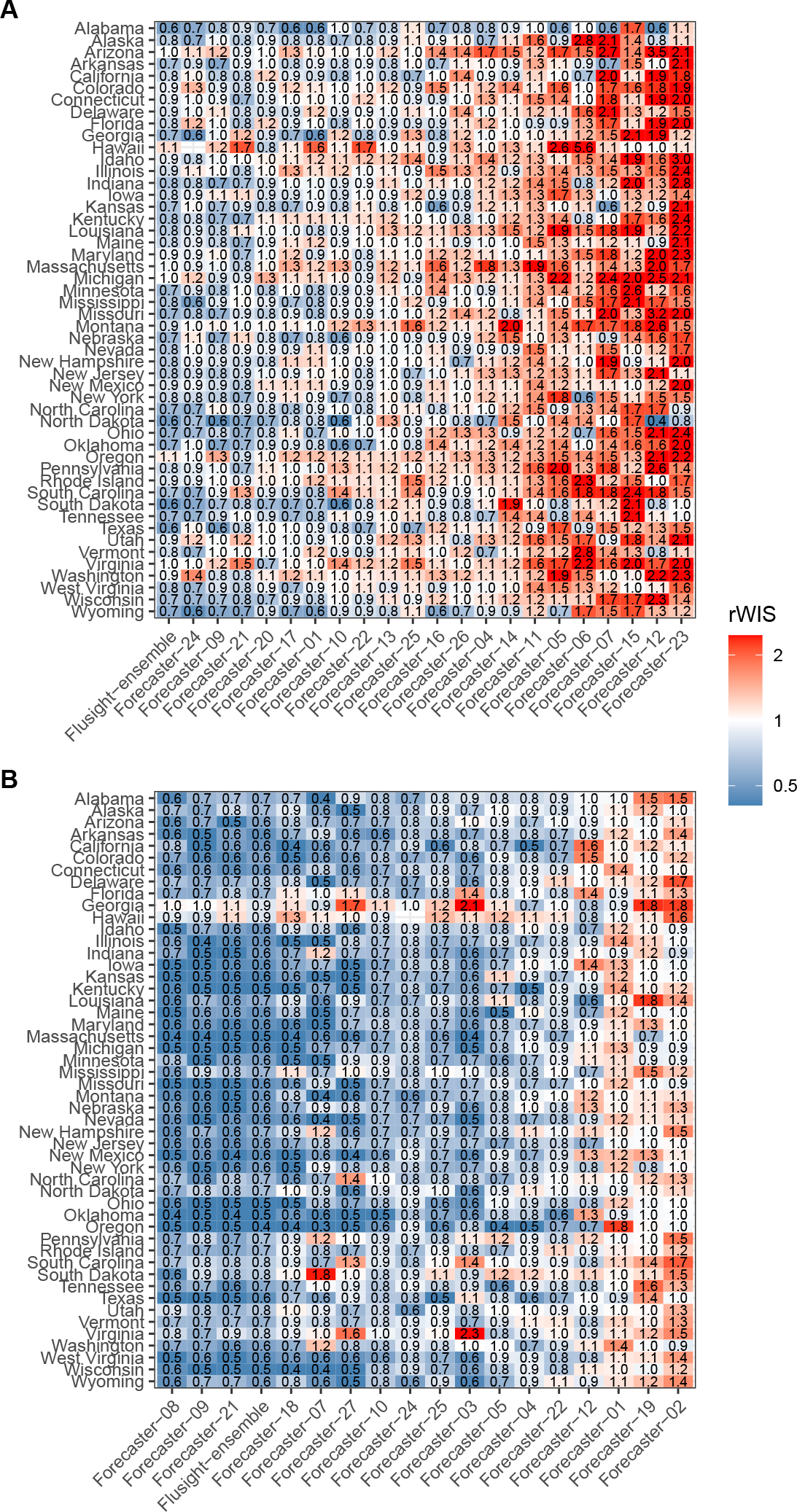
Heatmap of median relative weighted interval score (rWIS) across all forecasters eligible for evaluation within each season: (A) the 2021-22 season and (B) the 2022-23 season. Tiles are labeled to depict score and colored on a standardized scale within season. The forecasters are ordered by sum of rWIS across all locations. Locations are ordered alphabetically and only include the 50 states.

For the 2022-23 season (Fig. 2B) many more forecasters performed better than the baseline across most locations. However, forecasts for some locations, including Georgia and Hawaii, had higher rWIS (i.e., worse performance) across most of the forecasters. As with the 2021-22 season, some forecasters performed very well for certain locations but not as well for others. As an example, Forecaster-27 generally performed well across all locations except Georgia, Virginia, North Carolina, and South Carolina.

### 3.4 Performance across seasons

Forecasters submitting to both seasons did not all exhibit consistent performance across seasons. Fig. 3 shows the shift in ranks per median rWIS from 2021-22 to 2022-23. Several forecasters, including the FluSight ensemble, were consistently ranked high across both seasons. However, others shifted dramatically. For example, Forecaster-07 exhibited greatly improved performance in the 2022-23 season per the rWIS ranking. Likewise, relative to the other submissions Forecaster-01 performed much better in 2021-22 compared to 2022-23. Again, it is worth noting that the FluSight ensemble remained relatively constant. Appendix Fig. S1 provides a comparable rank change visualization using AE measures.

**Figure 3:**
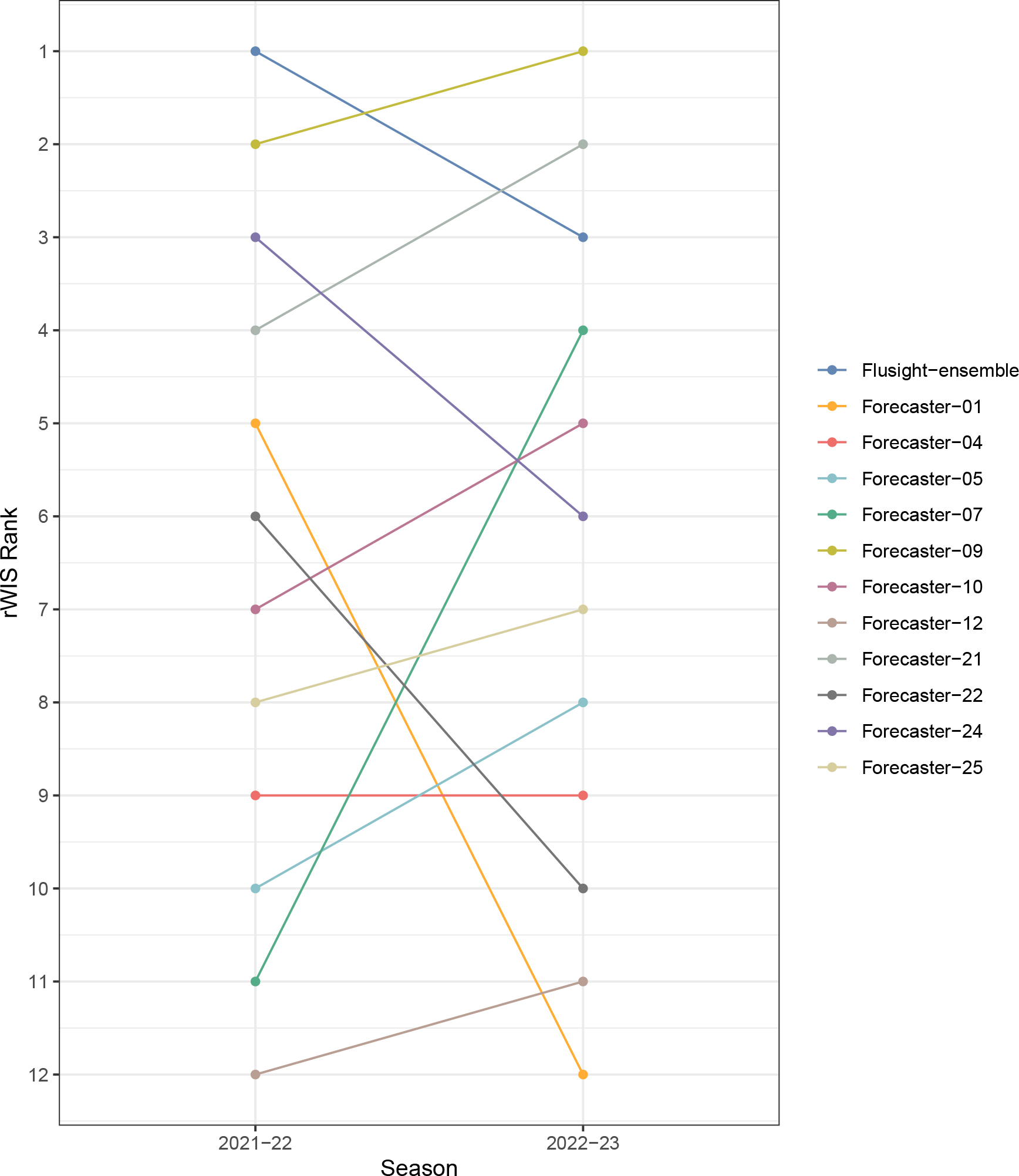
Forecasters ranked by median relative weighted interval score (rWIS) across seasons. The lower rank value indicates better performance (i.e., 1 is best). The line segments show direction of change (if any) in each forecaster rank in relation to others submitting to both seasons.

Fig. 4 provides a more granular depiction of performance at individual horizons and locations. The histogram shows the counts of individual rWIS rankings for submitted forecasts across all forecast weeks, horizons, and locations. Any given forecast ranked “1” performed the best relative to the other forecasters. The distributions of these ranks reinforce the finding that performance varies within season. For some forecasters we see different shapes of the rank distributions between 2021-22 and 2022-23, which communicates that the performance of methods used can vary across seasons as well. Appendix Fig. S2 provides a similar depiction of the distributions of the AE for forecasters within and between seasons.

**Figure 4:**
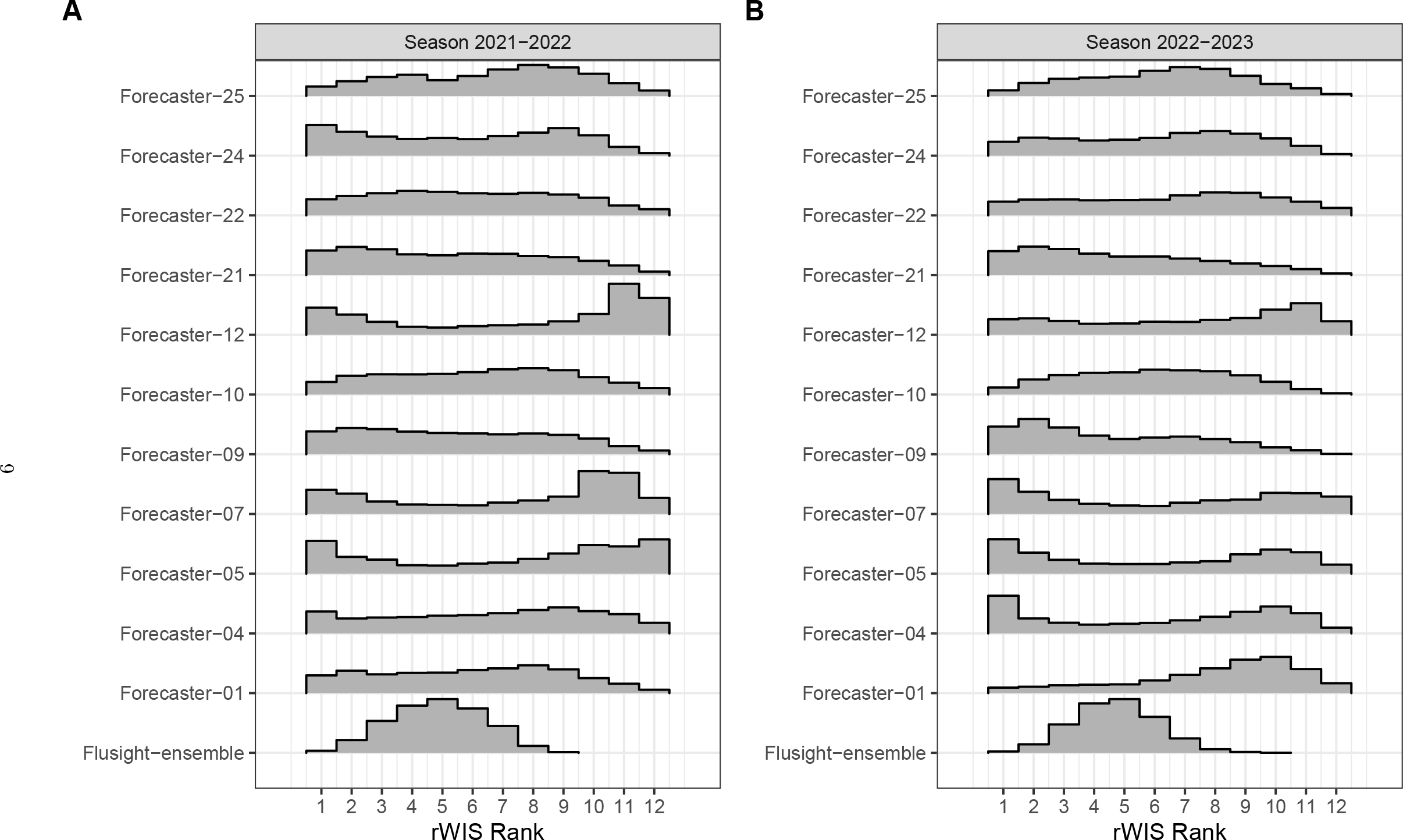
Counts of median relative weighted interval score (rWIS) ranks across all forecast weeks, locations, and horizons for (A) 2021-22 and (B) 2022-23 seasons. Scores are normalized to the baseline performance then ranked relative to one another. The lower rank value indicates better performance (i.e., 1 is best). Only forecasters submitting to both seasons are included.

Fig. 5 shows the absolute change between median WIS from 2021-22 and 2022-23. Note that for this analysis, the WIS is presented in place of the normalized rWIS to demonstrate magnitude change in performance improvement or degradation for each forecaster across seasons and location. For each forecaster, the WIS and AE are computed as median across forecast weeks and horizons within each forecaster and location in both seasons. The difference between the values (2022-23 median minus 2021-22 median) for every forecaster and location are visualized. Here, a larger negative value corresponds to greater improvement in performance in 2022-23 compared to 2021-22. It is worth noting that while the performance was generally higher relative to baseline for contributing forecasters in 2022-23 (Fig. 2B versus Fig. 2A), the heatmap of differences shows that the absolute change in median metrics for forecasters contributing to both seasons was generally modest. However, some forecasters (e.g., Forecaster-07 and Forecaster-12) improved WIS dramatically for many locations compared to the initial 2021-22 season. Appendix Fig. S3 displays the magnitude change in AE and corroborates the WIS finding.

**Figure 5:**
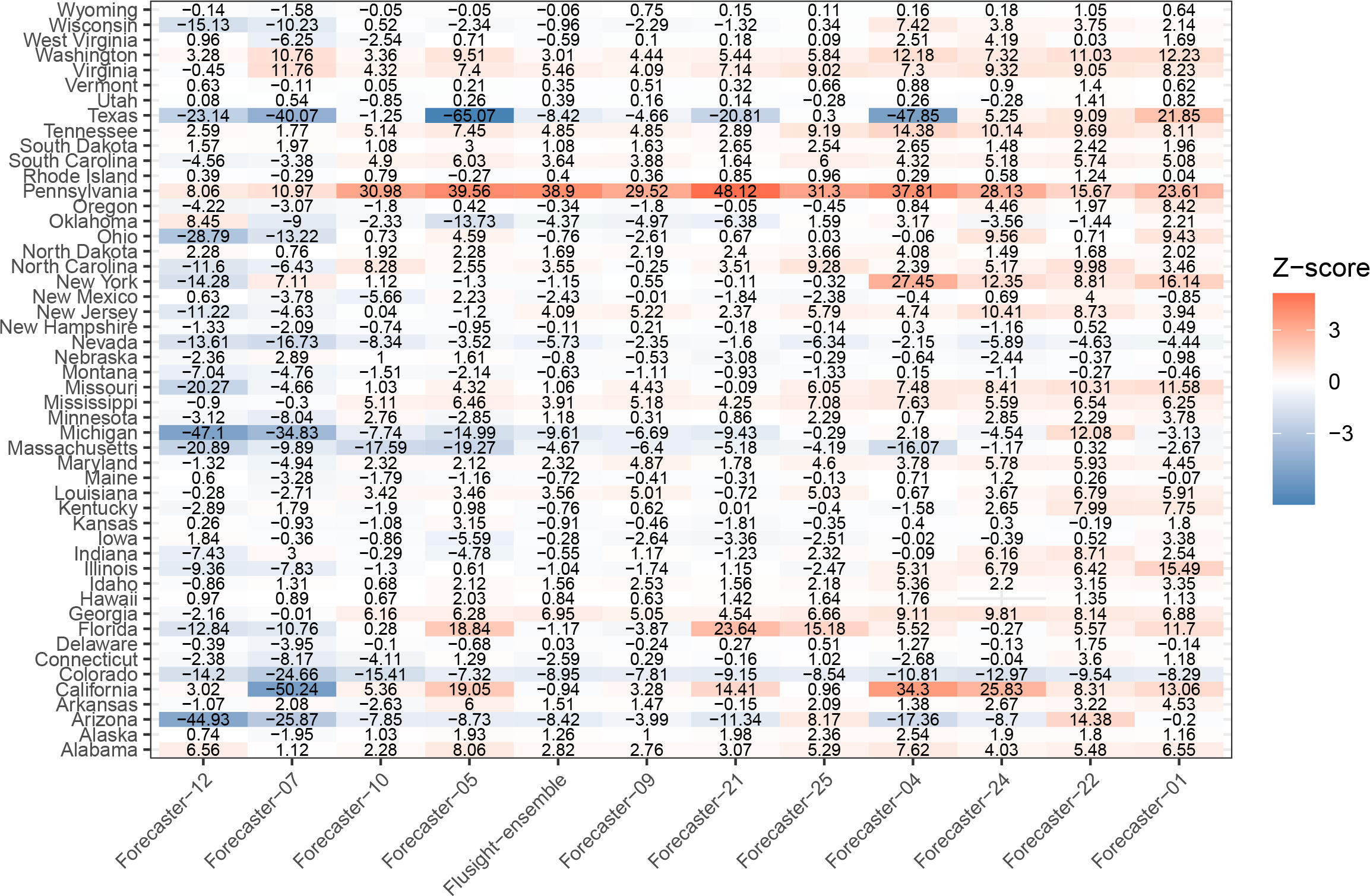
Difference in weighted interval score (WIS) between 2021-22 and 2022-23 seasons. The median value for the previous season is subtracted from the value for the more recent season, such that a negative difference indicates a drop in WIS (i.e., better performance). Forecasters are sorted by total sum of WIS difference across all locations. The heatmap is colored by standardized differences (Z-score).

## 4 Discussion

Our study provides several key insights regarding flu hospitalization forecasts submitted to FluSight across the 2021-22 and 2022-23 season: 1) the performance of forecasters generally varied by location; 2) contributing forecasters submitting in both 2021-22 and 2022-23 did not all perform the same across seasons; 3) the FluSight ensemble did consistently perform as one of the best forecasters in both seasons. FluSight invites submissions of forecasts at national, state, and territorial granularity. Our finding that forecaster performance varied across locations within each season affirms what we have observed through operational forecasting activities. Certain locations are more difficult to forecast, even for forecasters that otherwise perform well. It is worth noting that, the reverse scenario is also demonstrated in the results of this study. There were some forecasters that generally did not perform as well for most locations, but appeared to be especially well-suited for others and in fact outperformed all forecasters at these locations. This finding underscores the utility of the open ensemble approach, which inspires diverse methodological contributions. Some of those methods may not work well for all locations given location-specific dynamics of parameters and covariates used to train the models. However, if those methods do work well for specific locations then the ensemble could still benefit from the contributions. It is important to note that it is incumbent upon contributors to be conscientious with studying the locations for which their forecasters performed best/worst. Such review could lead to more targeted submissions that provide the most accurate information to the ensemble without contributing poorer performing forecasts.

The variability in performance of some of the forecasters contributing across seasons indicates that it is not necessarily safe to assume that a forecaster that works well for one season will work as well for the next. Likewise, a forecaster that performs poorly in a prior season may perform better later. This suggests that there may be challenges for forecasters who are evaluating forecast methods by looking at retrospective season performance. The finding also further demonstrates the utility of the consortium ensemble methods. In fact, while some other forecasters shifted in overall performance the FluSight ensemble was one of the best performers in both seasons.

Our study has several limitations. As described in the results, the 2021-22 and 2022-23 flu seasons were markedly different, with a much later peak in 2021-22 as compared to 2022-23. The 2021-22 season was also abbreviated due to limited hospitalization data reporting prior to January 2022. The difference in patterns of flu hospitalization incidence very likely impacted factors and covariates used to train and parameterize forecasting approaches. The different dynamics could also impact evaluations. For example, a season that was generally “easier” to forecast (e.g., long periods of stability in signal being forecast) might bias WIS towards lower values. While the different seasonal patterns could make it challenging to interpret absolute measures of performance our use of rWIS ranking should mitigate this effect. Across seasons, the rWIS may inherit some of the bias in different patterns observed. However, the shifts in ranks of rWIS is relatively robust in that all forecasters are compared to the baseline and relative to each other.

It is also worth noting that our assessment of forecast performance assumes that forecasters submitting to both seasons used the same methods. The FluSight coordinators require that forecasters include metadata describing methodology. We reviewed the GitHub history for the metadata file for all 11 contributing forecasters analyzed and did not find any major methodological changes noted. The consortium also asks that submitters use updated names (i.e., specifying a different forecaster altogether) if there are major changes to methods. With that said, it is still possible and perhaps likely that contributors would use lessons learned from the 2021-22 season and the intervening months to improve the forecasters moving into 2022-23. In this analysis, we cannot definitively say that methods used were identical within or between seasons. Furthermore, we know that while the FluSight ensemble methodology remained the same, it included forecasts from different constituent forecasters between 2021-22 and 2022-23.

## 5 Conclusion

We have studied the performance of flu hospitalization forecasters submitting near-term forecasts to FluSight during the 2021-22 and 2022-23 seasons. Generally, contributing forecasters performed better in some locations as compared to others in each season. If we assume that forecasters submitting to both seasons used the same methods in 2021-22 and 2022-23, then we can conclude that some methodology did not consistently perform the same. The FluSight ensemble forecast, however, did achieve some of the best overall performance in both 2021-22 and 2022-23. Collectively, these findings reinforce the need to continue to ensure that ensemble forecasting initiatives are adequately resourced with regular submissions from diverse methods. It is not reasonable to expect that a single independent forecasting approach will perform the best across all locations within a given season, nor when summarized and measured overall across seasons. The consortium ensemble, however, can benefit from information provided by independent forecasters to provide an accurate depiction of future disease activity and healthcare burden.

## Data Availability

All data produced in the present study are available upon reasonable request to the authors.

## 6 Acknowledgements

The work described in this manuscript would not have been possible without open and collaborative efforts from multiple entities. We acknowledge the following groups: the CDC for coordinating FluSight and providing guidance, interpretation, and dissemination of forecast data; the Council of State and Territorial Epidemiologists (CSTE) for establishing collaborative networks through which forecasting groups can interact with one another and public health stakeholders; the Reich Lab at the University of Massachusetts-Amherst for developing the Zoltar repository from which we retreived forecast data; all participating teams in the FluSight network for their sustained contributions, innovative techniques, and commitment to openness through operational forecasting activities.

This work was supported in part by a subaward to Signature Science, LLC from the CSTE via the CDC Cooperative Agreement No. NU38OT000297.

### Appendix

**Figure S1:**
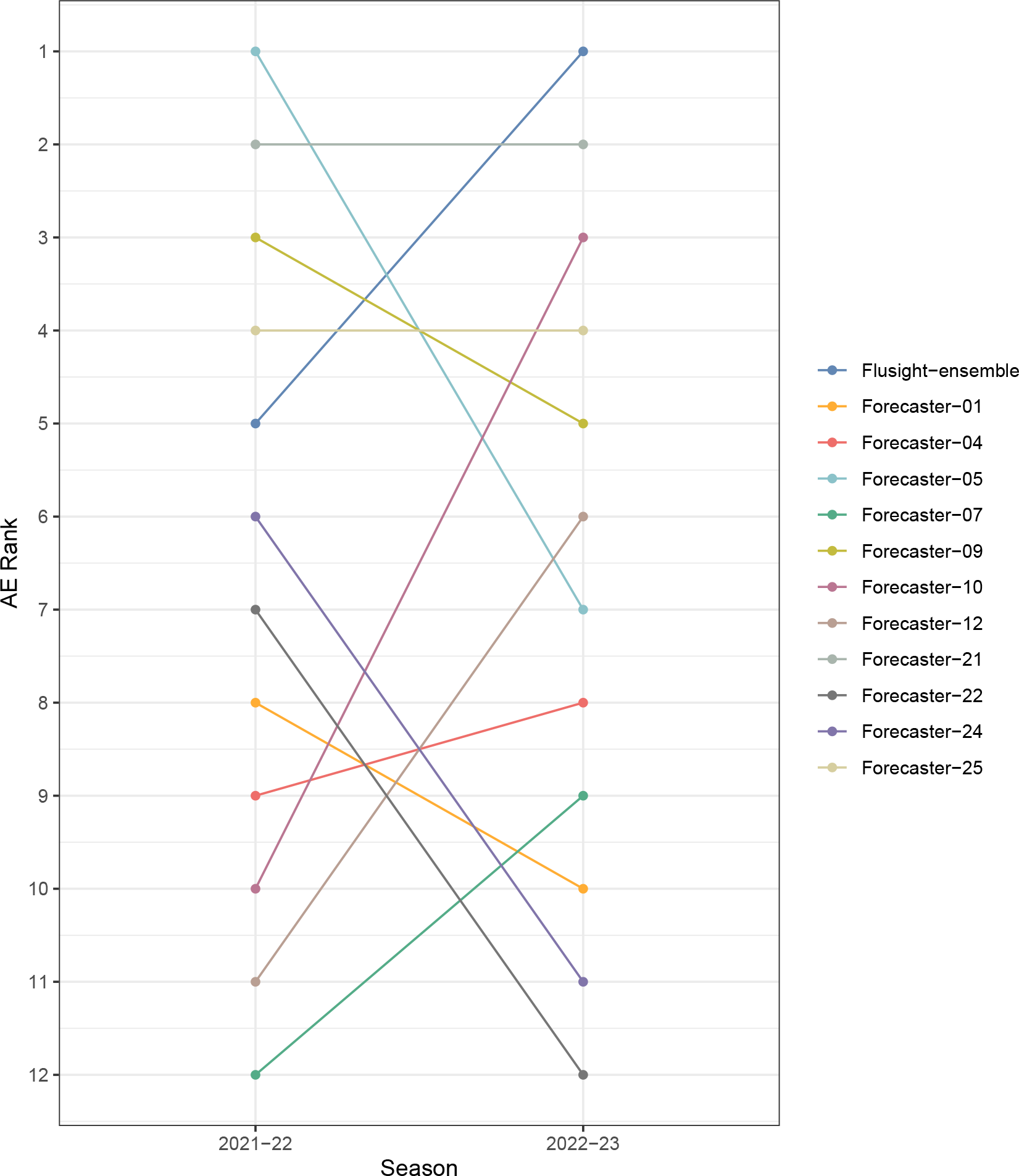
Forecasters ranked by median absolute error (AE) across seasons. The lower rank value indicates better performance (i.e., 1 is best). The line segments show direction of change (if any) in each forecaster rank in relation to others submitting to both seasons.

**Figure S2:**
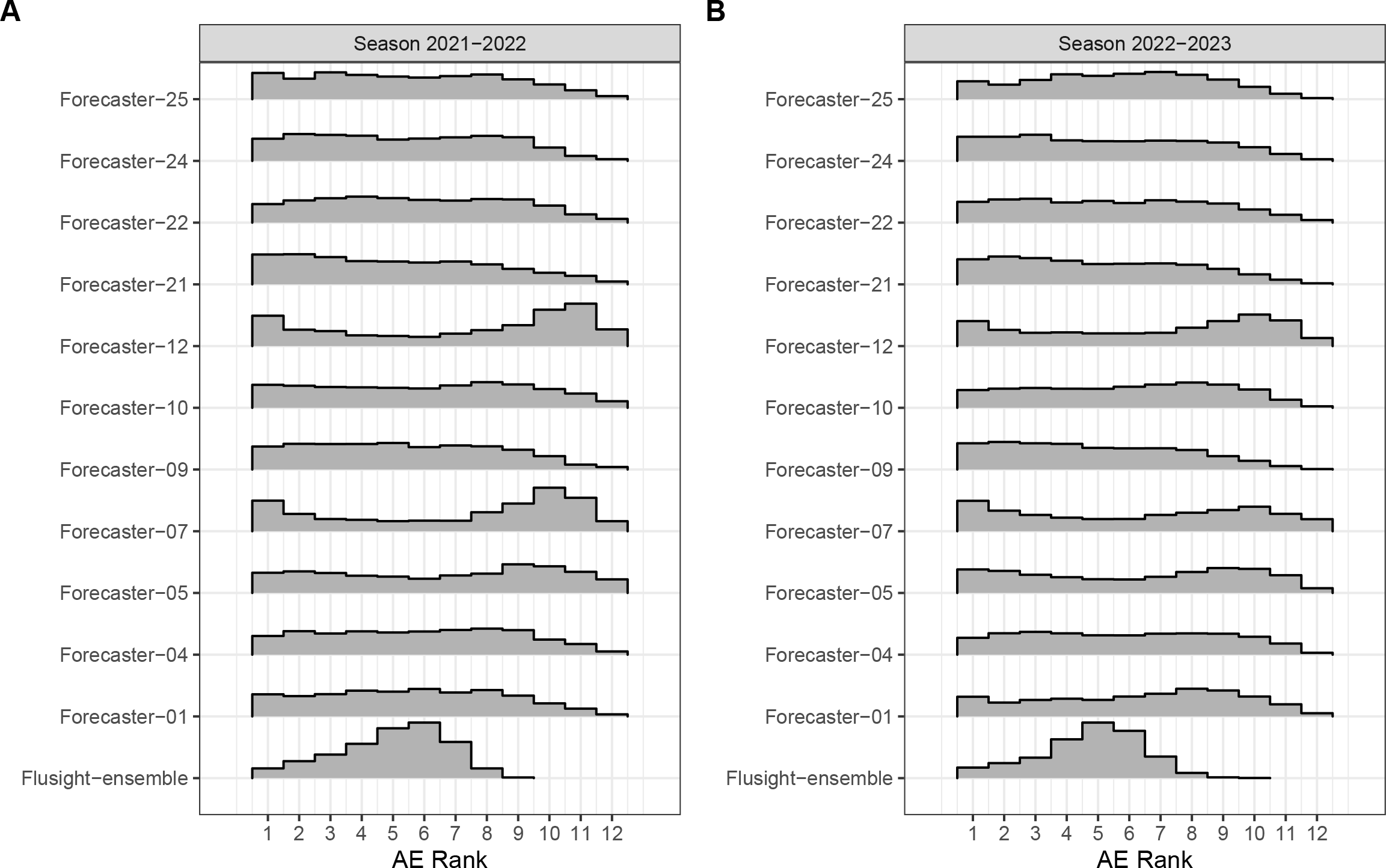
Counts of median absolute error (AE) ranks across all forecast weeks, locations, and horizons for (A) 2021-22 and (B) 2022-23 seasons. Forecasters are ranked relative to one another. The lower rank value indicates better performance (i.e., 1 is best). Only forecasters submitting to both seasons are included.

**Figure S3:**
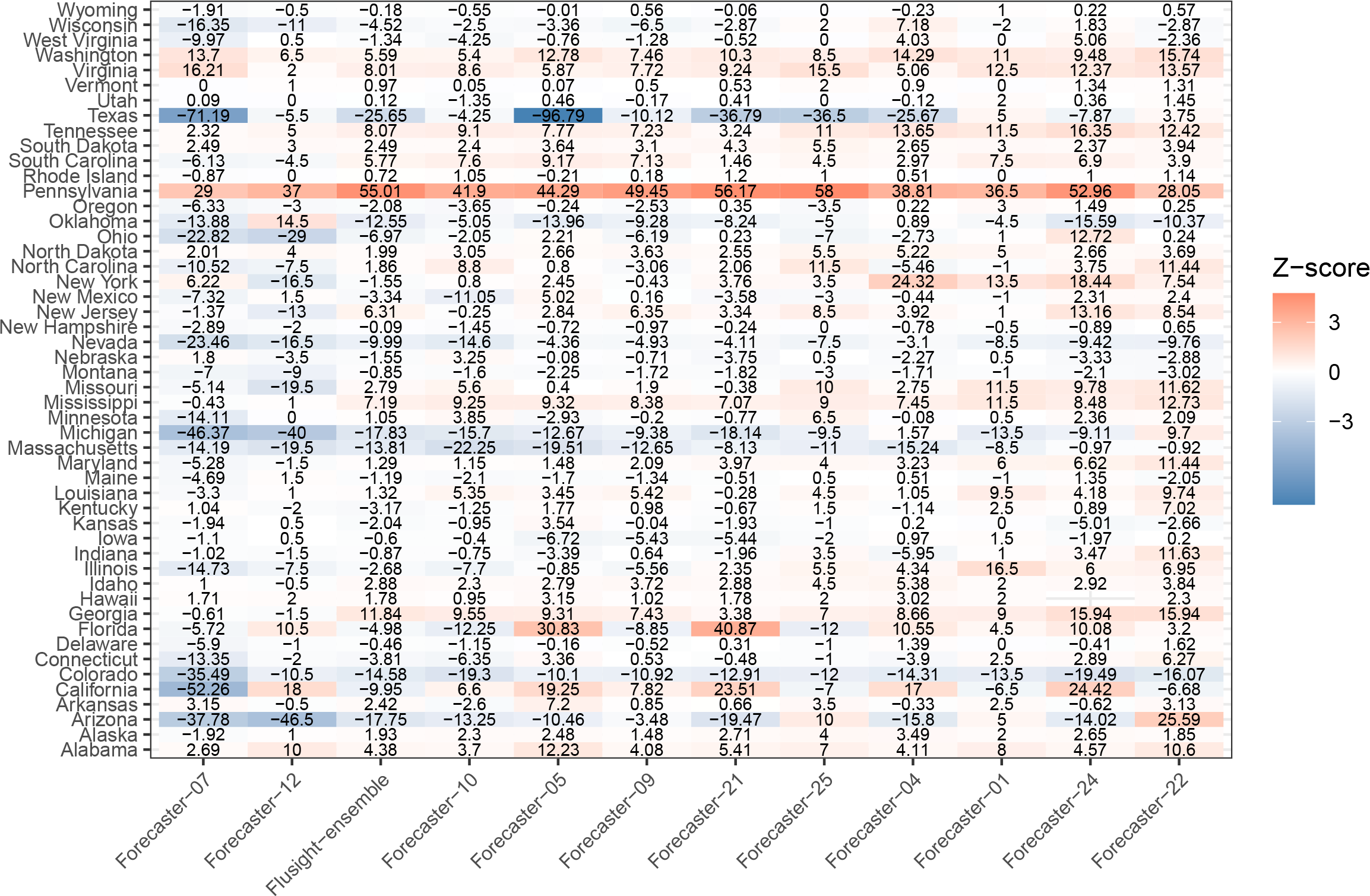
Difference in weighted interval score (AE) between 2021-22 and 2022-23 seasons by location. The median value for the previous season is subtracted from the value for the more recent season, such that a negative difference indicates a drop in AE (i.e., better performance). The heatmap is colored by standardized differences (Z-score).

## Footnotes

1 The forecast distribution is described via estimates for 23 quantiles: 0.01, 0.025, 0.05, 0.1, 0.15, 0.2, 0.25, 0.3, 0.35, 0.4, 0.45, 0.5, 0.55, 0.6, 0.65, 0.7, 0.75, 0.8, 0.85, 0.9, 0.95, 0.975, and 0.99.

2 To be eligible for inclusion in the ensemble, forecasts must be submitted ahead of the weekly deadline set by CDC. Forecasters designated as “primary”, “secondary”, or “proposed” will be included in ensemble.

